# Multinational, Calibrated, Non-Laboratory Prevalent Disease Prediction and Survival Modeling for Diabetes, CKD, and CVD

**DOI:** 10.64898/2025.12.10.25341981

**Authors:** Arthur Moreira Costa, Iris Badezet-Delory

## Abstract

Reliable non-laboratory tools for assessment of probability of prevalent disease (PPD) are essential for scalable prevention, yet existing models are typically specific to single diseases, require laboratory tests, and show no or limited calibration across PPD strata, limiting scaling and public health utilization. We developed and validated a unified, non-invasive machine-learning model for simultaneous prediction of diabetes, chronic kidney disease, and cardiovascular disease PPD non-invasive predictors. The model was trained on 2011–2016 National Health and Nutrition Examination Survey data (n=29,903) and evaluated on an independent 2017–2020 test set (n=15,559). It demonstrated moderate-to-strong discrimination (C-statistic=0.80–0.90), stable precision–recall performance, and moderate to strong calibration (slope>0.94). Validation in an independent Korean population showed no or minimal degradation in discrimination and calibration performance, though more extensive validation is warranted. Predicted PPD was associated with cause-specific mortality over up to 7 years of follow-up, consistent with a predictor of latent disease burden. Each 10-percentage-point increase in predicted PPD was associated with roughly a two-fold higher hazard of disease-specific death (HR 2.00–2.20). We conclude that this model has potential as scalable, low-burden screening/surveillance aid, but note that it is not intended as a diagnostic or prognostic tool.

## Introduction

Noncommunicable diseases (NCDs) remain the leading cause of global mortality, accounting for nearly three-quarters of non-pandemic-related deaths worldwide [1]. Their impact continues to intensify, with the Americas experiencing a 43% rise in NCD-related deaths since 2000 and exceeding six million deaths annually. In the United States, diabetes, chronic kidney disease (CKD), and cardiovascular disease (CVD) affect approximately 14.7%, 14%, and 9.9% of adults, respectively, with disproportionately higher burdens observed among older adults, Black individuals, and socioeconomically disadvantaged populations [1, 2, 3]. Because these chronic diseases share many modifiable factors and remain largely asymptomatic in early stages, scalable and low-burden approaches to predicting the probability of prevalent disease (PPD) are essential for effective prevention and public-health planning.

Despite considerable progress in PPD modeling, existing non-laboratory tools suffer from several constraints that limit their practical utility. Many rely on detailed self-reported dietary intake, which is known to exhibit substantial day-to-day variability, systematic underreporting, and high respondent burden [4, 5]. Other widely used tools lack calibration, restricting their use to coarse PPD categories rather than providing individualized, continuous PPD estimates necessary for informed decision-making and communication [6]. Moreover, most existing models are disease-specific, requiring individuals and public-health systems to administer separate tools for diabetes, CKD, and CVD despite substantial overlap in their non-laboratory predictors [7, 8]. Furthermore, most existing models are not externally validated and are thus limited to application in the country they were trained in. Together, these limitations reduce scalability, impede interpretability, and increase user burden, especially in population-level screening contexts.

To address these challenges, we evaluate whether a unified, low-burden, non-laboratory model can accurately predict PPD for diabetes, CKD, and CVD simultaneously. This model predicts PPD, not future incidence, and mortality associations reflect correlation with latent disease burden. Our models achieved discrimination comparable to established tools as well as calibration slopes close to 1 in both Korean and American populations. We also observe significant and consistent associations between model-predicted PPD and cause-specific mortality, supporting the role of our models as indicators of latent disease burden. The calibration of our models enables the use of continuous, individualized PPD estimates for three chronic diseases simultaneously, a significant improvement upon the typical coarse PPD estimates provided by the majority of the current approaches. The model estimates the probability that an individual already meets diagnostic criteria at the time of assessment using only non-laboratory predictors, but it does not estimate future risk or progression. It is intended as a scalable, low-burden screening and surveillance aid, not a diagnostic or prognostic tool.

## Results

### Population Description

Table S1 shows the characteristics of the weighted 2011-2020 National Health and Nutrition Examination Survey (NHANES) sample based on sex. There was an even distribution of sex, with participants averaging 37.74 ± 22.51 years old. Additionally, over 60 percent of the participants were identified as non-Hispanic whites, which is consistent with the demographic composition of the United States [9]. Furthermore, most participants had completed at least a high school education. Most women were non-smokers, but nearly half of men were smokers.

### Model Design

Figure 1 summarizes the modeling workflow, including data sources, preprocessing, model development, internal and external performance evaluation, and mortality association analysis. More detailed flow diagrams of model design and exclusion criteria are available in supplementary figures S1 and S2.

**Figure 1.**
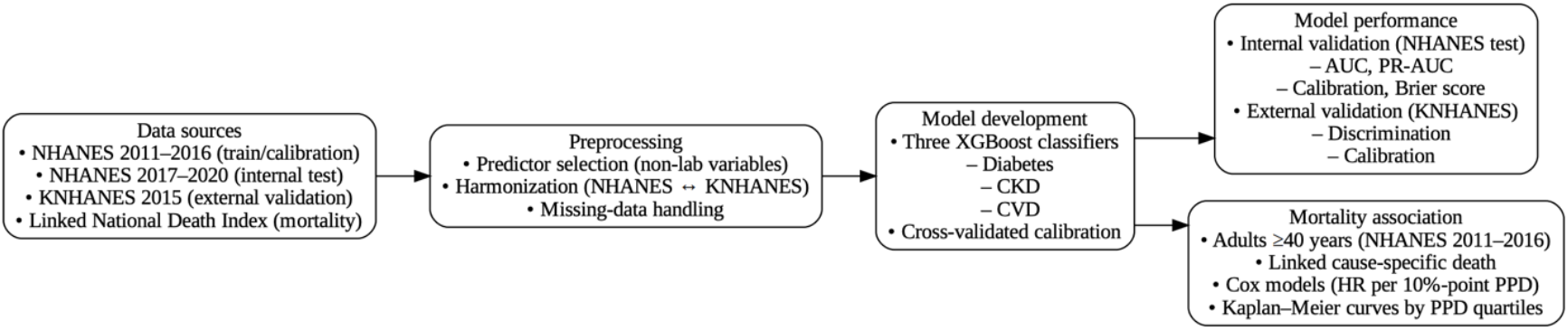
Model design pipeline.

### Model Discrimination

Figure 2 shows receiver operating characteristic (ROC) curves for diabetes, CKD, and CVD PPD prediction models and Area Under the ROC Curve (AUC), sensitivity, and specificity comparisons. All three models showed moderate to strong discrimination (AUC = 0.80-0.90) and sensitivity (0.79-0.89), with relatively weaker specificity (0.60-0.77).

**Figure 2.**
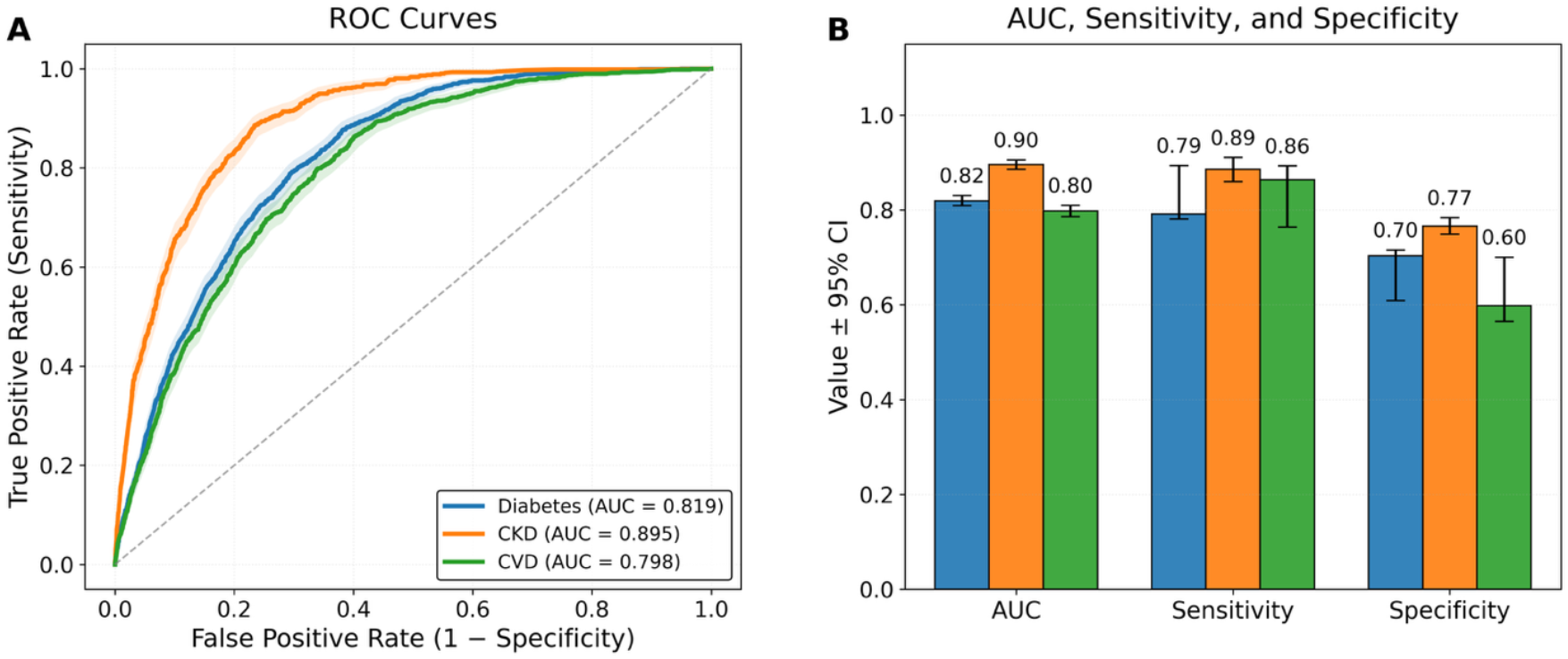
Model performance metrics. A) ROC curves for the diabetes, CKD, and CVD PPD-prediction models. B) Bar charts comparing AUC, sensitivity, and specificity between the models for each disease. CI bars and confidence ribbons represent the bootstrapped 95% CI with 1000 resamples of the test population with replacement.

### Precision-Recall and Calibration

Figure 3 displays precision–recall curves and calibration analyses after isotonic recalibration with 5-fold cross-validation, using training data for calibration. All of our models showed significantly higher PR-AUC than that of prevalence-based null models. Calibration curves indicated good alignment between predicted and observed PPD across deciles, with minimal over- or underprediction.

**Figure 3.**
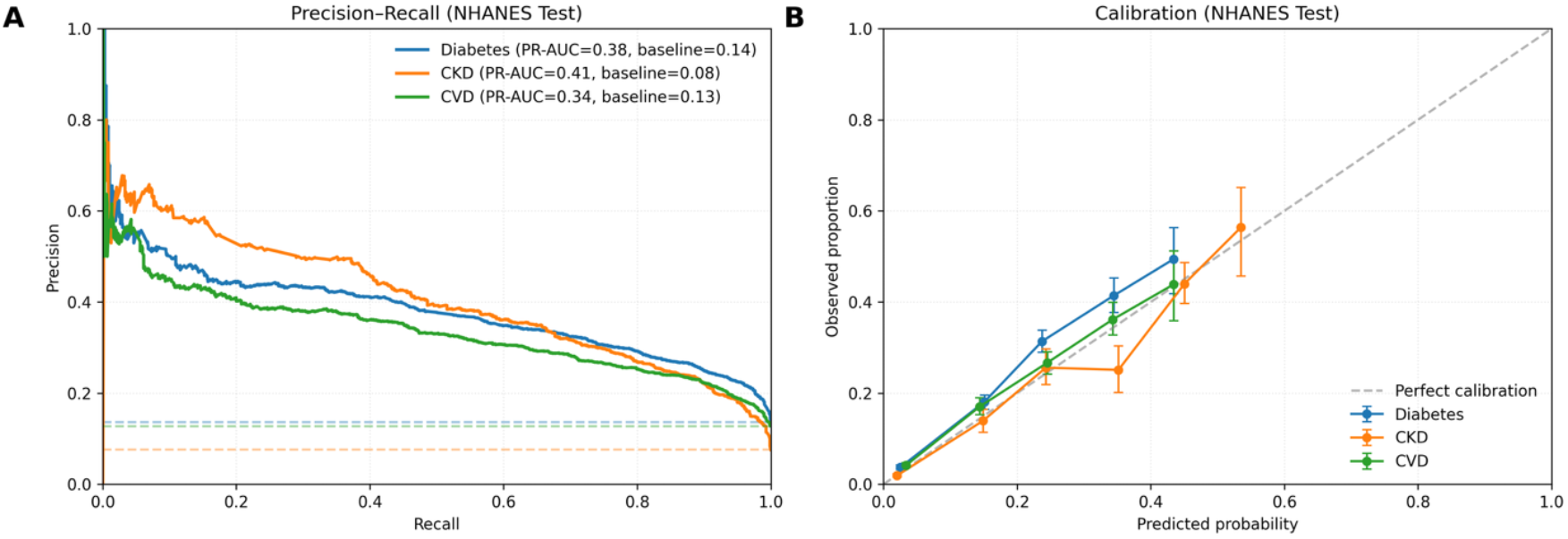
Precision-recall and calibration curves. A) PR-AUC is reported and PR curves are compared to prevalence baselines, show in dotted lines. B) Model calibration is compared to perfect calibration across decile predicted PPD bins. Bins with less than 50 individuals are omitted. Error bars represent 95% CIs.

### External Validation and Model Comparisons

Further evaluation of calibration performance was undertaken by quantitative comparisons of model Brier scores to prevalence-based controls (Table 1). All models achieved calibration slopes near 1 (slopes = 0.94-0.99). Diabetes had a particularly high intercept (0.288). All models were significantly better calibrated than null prevalence-based PPD estimators. Calibration performance in the 2015 Korean NHANES (KNHANES) cohort (Fig. S4; Table 1) showed strong calibration for diabetes and CVD (slopes > 0.99, |intercepts| < 0.02). To contextualize performance, we compared our models’ AUCs on NHANES and KNHANES with those reported for established non-laboratory models for diabetes, CKD, and CVD (Fig. 4).

**Table 1.** Cross-country comparison of discrimination, precision-recall, and calibration.

|  | U.S. |  |  | South Korea |  |  |
| --- | --- | --- | --- | --- | --- | --- |
| Metric | Diabetes | CKD | CVD | Diabetes | CKD | CVD |
| N | 9,908 | 9,475 | 9,231 | 5,824 | 7,380 | 6,459 |
| Prevalence (%) | 13.7 | 7.6 | 12.7 | 8.5 | 2.2 | 4.3 |
| AUC | 0.819 ± 0.010 | 0.895 ± 0.011 | 0.798 ± 0.012 | 0.768 ± 0.018 | 0.886 ± 0.019 | 0.868 ± 0.017 |
| PR AUC | 0.380 ± 0.025 | 0.405 ± 0.035 | 0.337 ± 0.026 | 0.183 ± 0.021 | 0.123 ± 0.029 | 0.188 ± 0.033 |
| Null Brier | 0.1179 | 0.07 | 0.1112 | 0.0779 | 0.0215 | 0.0408 |
| **Brier** | 0.1001 ± 0.0042 | 0.0544 ± 0.0032 | 0.0966 ± 0.0042 | 0.0723 ± 0.0052 | 0.0199 ± 0.0027 | 0.0365 ± 0.0033 |
| **ECE** | 0.0288 | 0.0061 | 0.0139 | <0.001 | <0.001 | <0.001 |
| **Calibration Slope** | 0.98 | 0.988 | 0.943 | 0.996 | 0.995 | 0.996 |
| **Calibration Intercept** | 0.288 | -0.097 | 0.058 | -0.008 | -0.014 | -0.008 |

**Figure 4.**
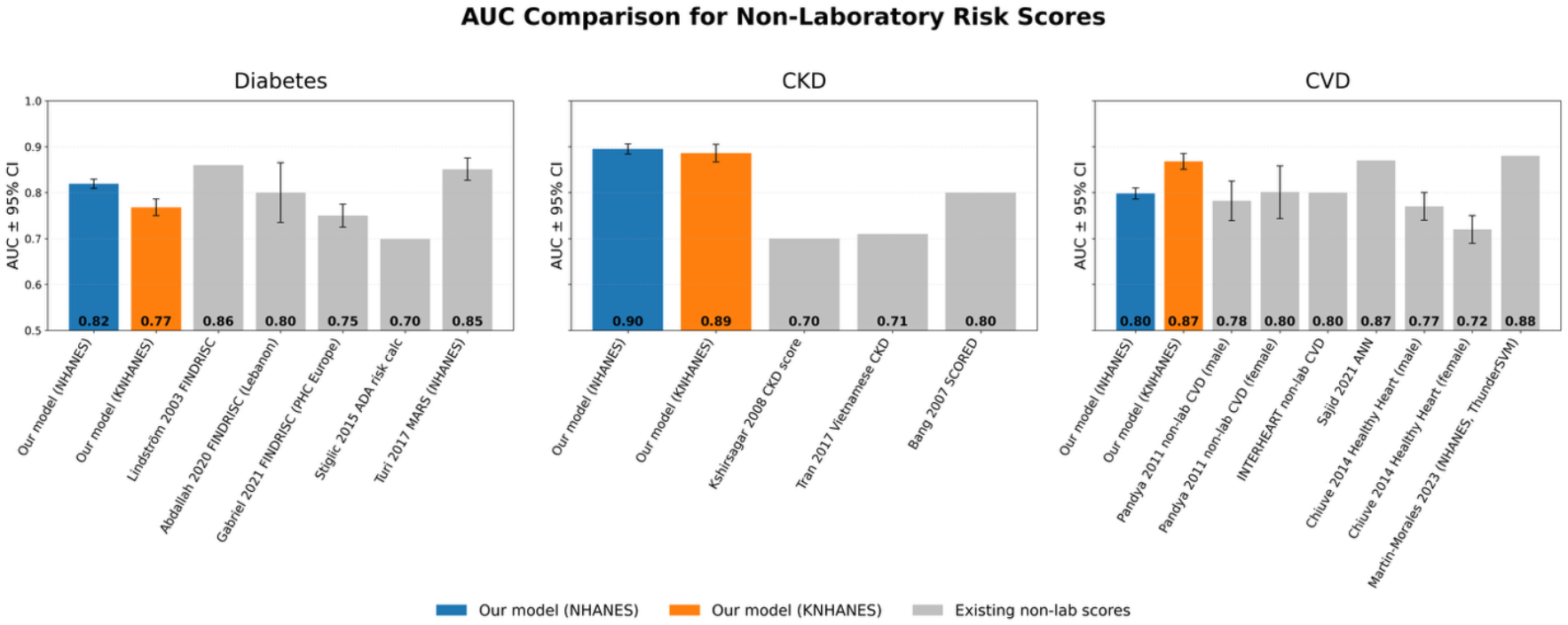
AUC comparisons between our model’s when tested in various countries, and other leading PPD prediction models [10-22]. Our models showed comparable calibration to leading predictive models when tested on both NHANES and KNHANES data. 95% CIs from external models are shown in models where they were reported.

### Survival analysis

Figure 5 shows validation of our predicted PPD against future cause-specific mortality. A 10-percentage-point increase in predicted diabetes, CKD, or CVD PPD was associated with approximately a two-fold higher hazard of disease-specific death (HR range: 2.00–2.20; p < 0.001 for all). When participants were stratified into quartiles of predicted prevalent CVD PPD, 7-year cumulative CVD mortality demonstrated clear monotonic separation (Fig. 5). Mortality increased from 0.0% (0.0–0.0) in Q1 to 1.2% (0.8–1.6) in Q4, representing more than a ten-fold gradient across quartiles (Table 2).

**Table 2.**
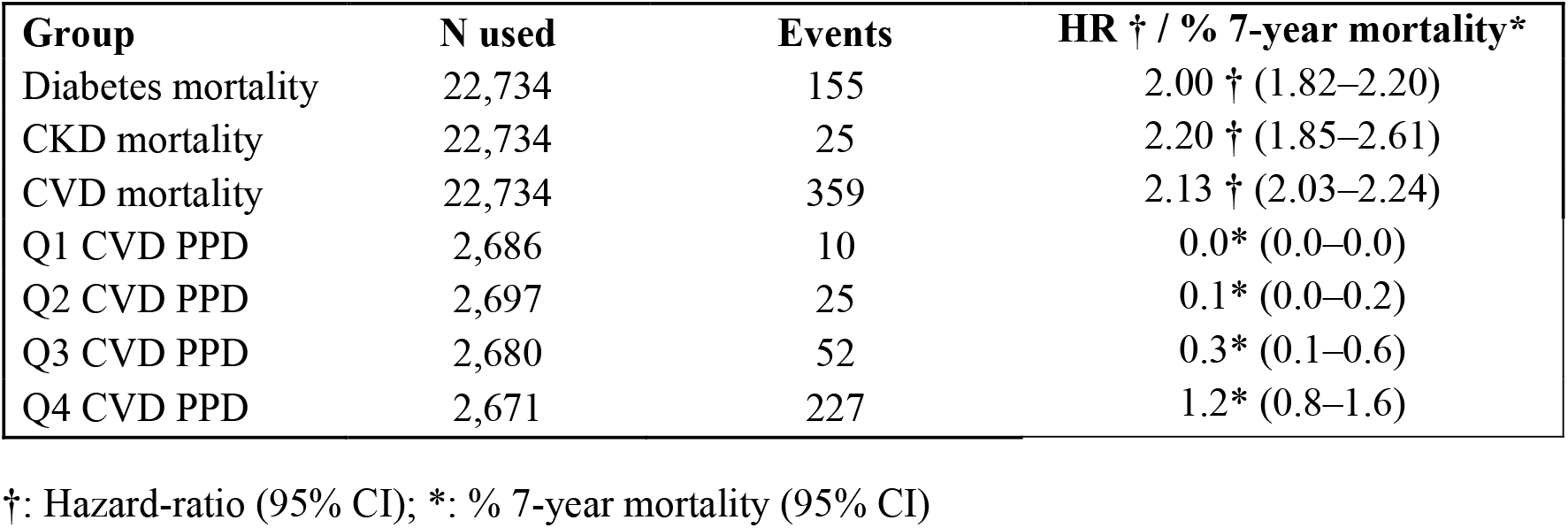
Hazard ratio and mortality by predicted-PPD quartiles.

**Figure 5.**
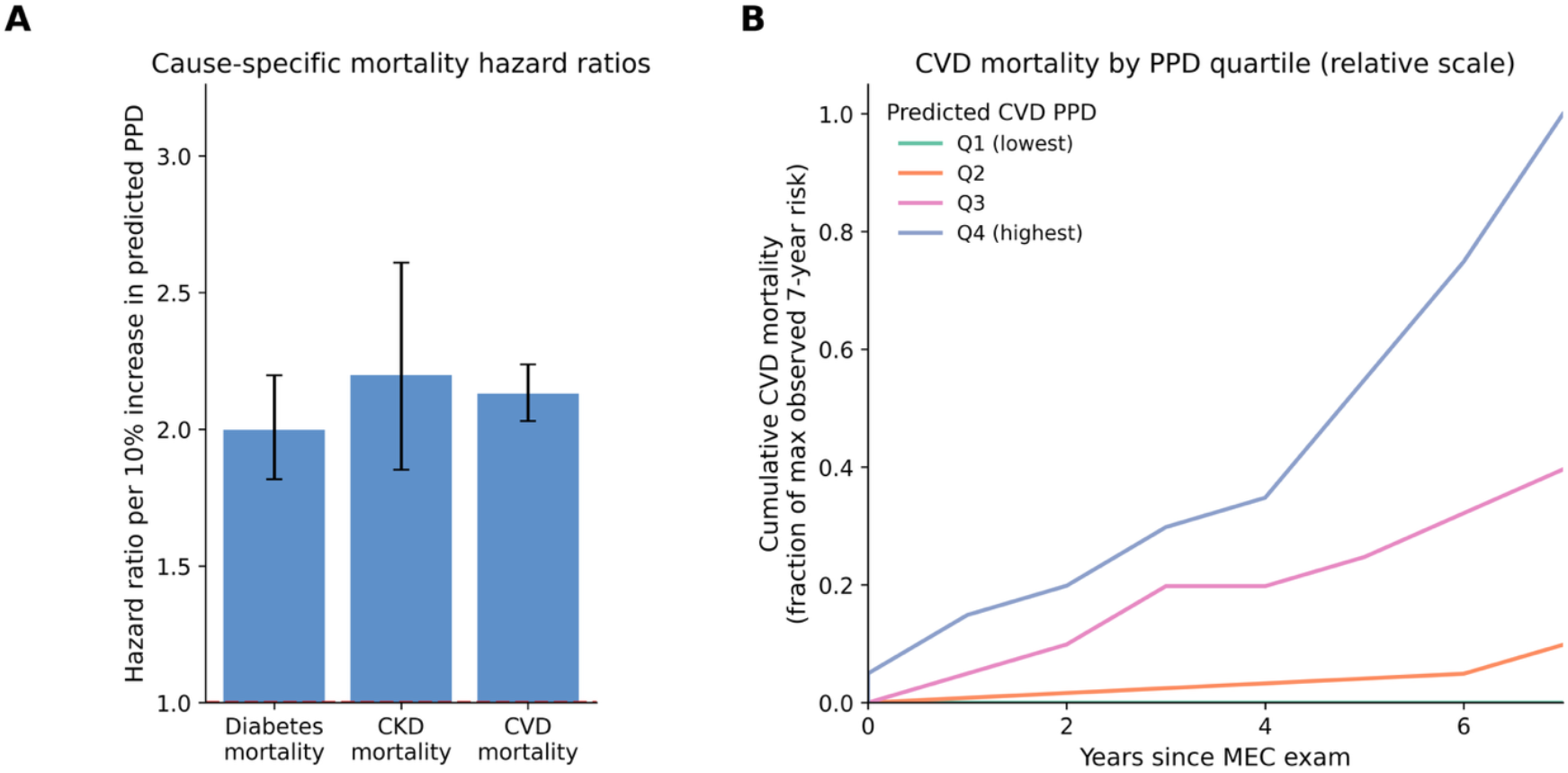
Validation of predicted PPD against cause-specific mortality and longitudinal CVD outcomes. **A)** Hazard ratios (HRs) for cause-specific mortality per 10-percentage-point increase in predicted PPD for diabetes, CKD, and CVD. Error bars indicate 95% CIs. **B)** Cumulative incidence of CVD mortality over 7 years, stratified by quartiles of predicted prevalent CVD PPD at baseline (Q1–Q4).

### Sensitivity Analyses

Sensitivity analyses showed that age, BMI, and waist circumference were the strongest predictors across diseases, consistent with partial dependence patterns. Feature-ablation experiments, including the removal or addition of dietary predictors, use of simpler tree ensembles, and alternative imputation strategies, did not materially change discrimination or calibration. Full results are provided in Supplementary Tables S2–S3 and Supplementary Figure S4.

## Discussion

We were able to create a model with comparable discrimination to current leading models, while also being well-calibrated, externally validated, and associated with future cause-specific mortality. It is also a single pipeline that enables simultaneous PPD prediction for three chronic diseases using a single set of general non-invasive predictors. When externally validating our model on 2015 KNHANES data without retraining, discrimination and calibration performance were comparable to the test set on 2017-2020 holdout test data from NHANES. Further comparison with established models indicates that our unified, three-disease model achieves discrimination on par with the leading noninvasive tools. Our CKD model showed particularly high discrimination, significantly higher than the leading models we analyzed. These results strengthen our claim that our tool can allow for decreased user burden by simultaneously screening for three diseases without losses in performance. Our survival analyses showed significant associations between predicted prevalent-disease probabilities and cause-specific mortality (Fig. 5; Table 2). These associations suggest that the model’s outputs do indeed correlate with underlying disease.

We then investigated how our lack of dietary predictors and our choice of boosted trees over simpler models were affecting model performance. We found that diet-aware models did not show improved calibration or discrimination. This finding is likely attributed to the low consistency and reliability of short-term dietary recall data collected by NHANES, rather than a physiological independence between diet and chronic disease PPD. We also found that simpler bagged tree models were unable to match the discrimination of our boosted tree models. Finally, we found near-identical performance of KNN and mean/mode imputation, which suggests that missing-data handling was not a major driver of model behavior, reducing concern that imputation introduced bias.

It is important to note, however, that all of our analyses are subject to significant limitations. Our population-level visualizations shown in figures S3 and S5 were generated using an unweighted population from our 2011-2016 NHANES training data. This population is not representative of the true US population and should not be extrapolated to make conclusions about disease-burdens on a full population. Additionally, although our model was ported to a Korean population (KNHANES), portability to lower-middle-income countries was not analyzed and should be addressed in future work. Another limitation is that our analyses are all association-based, and causal inferences cannot be drawn from our model. Similarly, our model does not indicate longitudinal or incident risk. Because the model is trained on cross-sectional data, predicted probabilities should not be interpreted as forecasts of future incidence or as estimates of the effects of behavioral or clinical interventions. Future work should assess this framework in prospective cohorts, incorporate repeated measurements, or evaluate whether longitudinal changes in predicted probabilities are associated with subsequent differences in long-term outcomes.

## Materials and Methods

### Study population and weighting

We used NHANES 2011–2020 data to train and test our model [24]. NHANES 2011-2016 data was used for training and calibration (n = 29903), and 2017-2020 data was used for testing (n = 15,559). Sampling weights were applied for descriptive statistics but not for machine learning model development. We did not apply survey weights during model training because our goal was to learn the individual-level conditional probability P(Y=1|X), not to estimate population-level prevalence. Moreover, weighting increases population representativeness but doesn’t improve predictive accuracy can degrade individual-level risk estimation. Furthermore, NHANES weights can distort model fitting by over-emphasizing subgroups that make up larger proportions of the US population. Unweighted training therefore provides a cleaner estimate of the covariate-outcome relationship, while absolute risk alignment is handled separately through calibration and external validation. Weights for descriptive statistics were calculated according to NHANES guidelines using mobile examination center weights [24]. Weights were normalized to a mean value of 1.00, which is a standard procedure used in other NHANES-based analyses [e.g. 25, 26].

### Predictors and outcome

Standard predictors included demographic variables (age, sex, income-to-poverty ratio), anthropometric measures (body mass index (BMI), waist circumference), and physiologic measures (blood pressure, heart rate). Dietary predictors included dietary sugar, saturated fat, carbohydrate, fiber, polyunsaturated fat, mineral, vitamin, caffeine, theobromine, alcohol, and caloric intake. Diabetes status was defined as glycohemoglobin (HbA1c) ≥ 6.5%, consistent with the recommendation of the American Diabetes Association [27]. CKD status was determined according to KDIGO 2012 criteria as eGFR <60 mL/min/1.73m^2^, using the CKD-EPI equation [28]. CVD status was designated as a self-reported history of coronary heart disease, congestive heart failure, angina pectoris, heart attack, or stroke. For external validation on the KNHANES data, CVD was defined as a positive self-reported history of myocardial infarction or angina. Missing predictor values were imputed using training-set means for continuous variables, and modes for categorical variables. Participants missing HbA1c were excluded from diabetes analyses. Those missing any variables needed to compute eGFR (most commonly serum creatinine) were excluded from CKD analyses. Those missing any variables used to define CVD status were excluded from CVD analyses. Predictors in external validation were manually converted to NHANES-coded variables. A variable dictionary and missingness table are shown in tables S4 and S5.

### Model training and evaluation

An initial model screening, trained on NHANES 2011–2016 data and tested on independent 2017–2020 data, was conducted. Models tested included logistic regression, regularized logistic regression, naïve Bayes, k-nearest neighbors, decision trees, random forests, bagged trees, boosted trees (AdaBoost and gradient boosting), and support vector machines with linear and nonlinear kernels. Across these models, tree-based classifiers showed the lowest classification error. The gradient boosting framework XGboost was selected for further development because it achieved the highest discrimination while offering stable performance across resamples, favorable calibration behavior with minimal hyperparameter tuning, robustness to moderate shifts in predictor distributions between NHANES and KNHANES, and clean integration with permutation-based interpretability and partial-dependence analyses. Model hyperparameters are described in the supplementary information, with key parameters listed in table S6.

Model performance was evaluated using sensitivity, specificity, area under the Receiver Operating Characteristic (ROC) curve (AUC), and Brier score. To investigate whether we could exclude dietary variables without sacrificing predictive performance, four models were explored for each disease: a 48-hour-recall diet model (which averages day 1 and day 2 recalls into mean intake variables), a 24-hour-recall diet model, a standard predictor model, and a standard predictor + 48-hour dietary recall model. All model thresholds were chosen to maximize Youden’s J (i.e. sensitivity + specificity -1). To account for total energy intake, we created energy-adjusted dietary variables (nutrient intake per 1,000 kcal) and repeated diet-only models with these adjusted features. AUC was calculated for each model and was used to compare discrimination performance between models.

In addition to ROC analysis, we evaluated model performance using precision–recall (PR) curves and area under the PR curve (PR-AUC), which is more informative under class imbalance. Calibration (reliability) curves were generated using decile bins to assess agreement between predicted and observed PPD. We also computed bootstrap 95% confidence intervals (CIs) for AUC using bootstrap resampling. Null Brier scores for each chronic disease were calculated by creating hypothetical models that assign each individual in the test population a PPD estimate equal to the prevalence in the studied population/subpopulation.

Our external validation was done by curating the 2015 KNHANES dataset such that variable labels and values were matched to NHANES data used to train the models, except for CVD, which could only be partially matched, as discussed above [29]. We then compared our model to other external models, selected via a literature search for non-invasive diabetes, CKD, and CVD PPD models, prioritizing varied recent and well-established models (e.g. FINDRISC, INTERHEART).

### Partial-dependence analyses

To examine how predicted probabilities vary with individual predictors, we computed partial-dependence sensitivity analyses using the observed NHANES training sample. For each grid value, we replaced the predictor value for all individuals in the training set while leaving all other predictors unchanged, passed the modified dataset through the trained model, and calculated the mean predicted probability for each disease. This yields a one-dimensional partial-dependence curve that summarizes how the model’s predictions vary with that predictor, averaged over the joint distribution of all other predictors in the training sample.

### Cox Models, Discrimination, and Survival Across Predicted PPD Quartiles

Cause-specific mortality was obtained from the 2019 NHANES-linked National Death Index files. Follow-up time was calculated from the MEC examination using PERMTH_INT (months) and converted to years. Participants outside the 2011–2016 survey cycles or missing survival time, cause-of-death indicators, or predicted-PPD estimates were excluded. To ensure meaningful exposure windows for chronic-disease mortality and to match established NHANES cardiovascular mortality analyses, we restricted the survival analyses to adults aged ≥40 years. Event indicators were defined for diabetes, CKD, and cardiovascular mortality using the underlying-cause classification provided in the linked files.

For each mortality endpoint, we fit pooled Cox proportional hazards models across all eligible NHANES cycles, including cohort indicators and scaling predicted PPDs so hazard ratios represent the effect of a 10-percentage-point increase in model-predicted PPD. Cox coefficients were estimated using a numerically stable Breslow partial-likelihood implementation with ridge penalization, which improves convergence for sparse endpoints (e.g., CKD mortality). To evaluate PPD stratification, predicted CVD PPD was divided into quartiles, and 7-year cumulative CVD mortality was estimated within each quartile using Kaplan–Meier curves and bootstrap-derived confidence intervals. Similar analyses were not conducted for diabetes or CVD due to insufficient cause-specific mortality.

## Abbreviations and Acronyms

PPD: probability of prevalent disease
NCD: noncommunicable disease
DM: diabetes mellitus
CVD: cardiovascular disease
CKD: chronic kidney disease
NHANES: National Health and Nutrition Examination Survey
CI: confidence interval
AUC: area under curve
ROC: receiver operating characteristic
BMI: body mass index
HbA1c: glycohemoglobin
KDIGO: Kidney Disease Improving Global Outcomes
PR: precision-recall
PR-AUC: area under the precision-recall curve
eGFR: estimated glomerular filtration rate
CDC: Centers for Disease Control
PDP: partial dependence plot

## Acknowledgments

The authors thank Gabriela Gomez, Dr. Ismael Haddadian, and Dr. Sandra C. Fuchs for helpful feedback.

## Author Contributions

A.M.C completed the conceptualization, methodology, and formal analysis. Investigation and data curation was completed by A.M.C and I.B.D. The first draft of the manuscript was written and edited by A.M.C and I.B.D. All authors have read and agreed to the published version of the manuscript.

## Ethics statement

NHANES and KNHANES are publicly available, deidentified national health surveys that obtain informed consent from all participants and are approved by their respective institutional review boards. This secondary analysis of fully anonymized public data was exempt from institutional review board review.

## Informed Consent Statement

All participants in NHANES and KNHANES provided written informed consent.

## Data Availability

All data used in this study are publicly accessible. NHANES 2011–2020 data can be obtained from the U.S. Centers for Disease Control and Prevention (https://www.cdc.gov/nchs/nhanes). KNHANES 2015 microdata are available from the Korea Disease Control and Prevention Agency via the KNHANES website (https://knhanes.kdca.go.kr) following free online registration; the authors do not redistribute these files. Mortality data were obtained from the NHANES-linked National Death Index. No proprietary or restricted-access datasets were used.

## Code availability

All analysis code used in this study are available in repositories linked in the supplementary information.

## References

1. Centers for Disease Control and Prevention. Diabetes: Data & Research. Atlanta, GA: CDC. (accessed 4 Sept 2025).

2. Centers for Disease Control and Prevention. Chronic Kidney Disease: Data & Research (Fast Stats). Atlanta, GA: CDC. (accessed 4 Sept 2025).

3. Martin SS, et al. Heart Disease and Stroke Statistics—2024 Update: A Report From the American Heart Association. Circulation. 2024;149:e347–e913.

4. Poslusna K, et al. Misreporting of Energy and Micronutrient Intake Estimated by Food Records and 24 h Recalls, Control and Adjustment Methods in Practice. Br J Nutr. 2009;101(S2):S73–S85.

5. Ahluwalia N, et al. Update on NHANES Dietary Data: Focus on Collection, Release, Analytical Considerations, and Uses to Inform Public Policy. Adv Nutr. 2016;7:121–134.

6. Van Calster B, et al. Calibration: The Achilles Heel of Predictive Analytics. BMC Med. 2019;17:230.

7. Lindström J, Tuomilehto J. The Finnish Diabetes Risk Score (FINDRISC): A Valid Tool to Estimate the Risk of Type 2 Diabetes. Diabetes Care. 2003;26(3):725–731.

8. D’Agostino RB Sr, et al. General Cardiovascular Risk Profile for Use in Primary Care: The Framingham Heart Study. Circulation. 2008;117:743–753.

9. CDC/NCHS. Health, United States—Sources and Definitions: National Health and Nutrition Examination Survey (NHANES). (accessed 4 Sept 2025).

10. Zhou X, Qiao Q, Ji L, et al. Non-laboratory-based risk assessment algorithm for prevalent type 2 diabetes developed on a nation-wide diabetes survey. Diabetes Care. 2013;36(12):3944–3952.

11. Abdallah L, Naja F, Choufani J, et al. Validation of the Finnish Diabetes Risk Score (FINDRISC) in the Lebanese population. Prim Care Diabetes. 2020;14(3):271–277.

12. Mugume I, Kibirige D, Ssebunya R, et al. Performance of the Finnish Diabetes Risk Score in detecting prevalent type 2 diabetes and dysglycaemia in a rural Ugandan population. PLOS One. 2023;18(11):e0276858.

13. Ha KH, Kim DJ. Development and validation of the Korean Diabetes Risk Score: a prospective study. Diabetologia. 2018;61(4):949–958.

14. Zhou X, Li W, Wong CKH, et al. Simple non-laboratory and laboratory-based risk assessment algorithms for prevalent diabetes in a Chinese population. J Diabetes Investig. 2016;7(5):657–666.

15. Bang H, Vupputuri S, Shoham DA, et al. SCreening for Occult REnal Disease (SCORED): a simple prediction model for chronic kidney disease. Arch Intern Med. 2007;167(4):374–381.

16. O’Seaghdha CM, Lyass A, Massaro JM, et al. A risk score for chronic kidney disease in the general population. Am J Med. 2012;125(3):270–277.

17. Tangri N, Stevens LA, Griffith J, et al. A predictive model for progression of chronic kidney disease to kidney failure. JAMA. 2011;305(15):1553–1559.

18. Gaziano TA, Young CR, Fitzmaurice G, Atwood S, Gaziano JM. Laboratory-based versus non-laboratory-based method for assessment of cardiovascular disease risk: the NHANES I Follow-up Study cohort. Lancet. 2008;371(9616):923–931.

19. Pandya A, Weinstein MC, Gaziano TA. A comparative assessment of non-laboratory-based versus commonly used laboratory-based cardiovascular disease risk scores in the NHANES III population. PLOS One. 2011;6(5):e20416.

20. McGorrian C, Yusuf S, Islam S, et al. Estimating modifiable coronary heart disease risk in multiple regions of the world: the INTERHEART Modifiable Risk Score. Eur Heart J. 2011;32(5):581–589.

21. Kaptoge S, Pennells L, De Bacquer D, et al. World Health Organization cardiovascular disease risk charts: revised models to estimate risk in 21 global regions. Lancet Glob Health. 2019;7(10):e1332–e1345.

22. Rezaei F, Yadegarfar G, Gohari K, et al. Agreement between laboratory-based and nonlaboratory-based Framingham risk scores in a large Middle Eastern cohort. Sci Rep. 2021;11:10687.

23. Jia, G.; Aroor, A.R.; Sowers, J.R. Arterial Stiffness: A Nexus between Cardiac and Renal Disease. Cardiorenal Med. 2014, 4, 60–71. 10.1159/000360867.

24. University of Texas at Arlington. National Health and Nutrition Examination Survey (NHANES)—Big Data for Epidemiology. Available online: https://uta.pressbooks.pub/bigdataforepidemiology/chapter/chapter10-nhanes/ (accessed on 4 September 2025).

25. Costa, A.M.; Sias, R.J.; Fuchs, S.C. Effect of Whole Blood Dietary Mineral Concentrations on Erythrocytes: Selenium, Manganese, and Chromium: NHANES Data. Nutrients 2024, 16, 3653. 10.3390/nu16213653.

26. Chen, J.; Kan, M.; Ratnasekera, P.; Deol, L.K.; Thakkar, V.; Davison, K.M. Blood Chromium Levels and Their Association with Cardiovascular Diseases, Diabetes, and Depression: NHANES 2015–2016. Nutrients 2022, 14, 2687. 10.3390/nu14132687.

27. American Diabetes Association. Diabetes Diagnosis. Available online: https://diabetes.org/about-diabetes/diagnosis (accessed on 4 September 2025).

28. World Health Organization. Noncommunicable Diseases – Fact Sheet. Geneva: WHO. (accessed 4 Sept 2025).

29. Kweon, Sanghui et al. “Data Resource Profile: The Korea National Health and Nutrition Examination Survey (KNHANES)” vol. 43, no. 1, 2014

