## Supplemental Information for "Multinational, Calibrated, Non-Laboratory Prevalent Disease Prediction and Survival Modeling for Diabetes, CKD, and CVD"

### Supplementary Information – Table of Contents

#### Supplementary Figures


- Figure S1. Full modeling pipeline diagram
- Figure S2. Flow of participants through the NHANES analytic pipeline
- Figure S3. Feature importance analyses (tree-based and permutation-based)
- Figure S4. Precision–recall and calibration curves for KNHANES
- Figure S5. Partial dependence plots for diabetes, CKD, and CVD models


#### Supplementary Tables


- Table S1. Demographic characteristics of the weighted sample
- Table S2. Model performance with and without dietary predictors
- Table S3. Naïve vs. final model comparison
- Table S4. Variable missingness summary
- Table S5. Variable dictionary and harmonization mapping
- Table S6. XGBoost model hyperparameters


Trained models/Datasets/Online-tool code:
https://github.com/risk-prediction-research/NSR_models

Figure/Table creation code:
https://colab.research.google.com/drive/1G7M2g-yc9sv2OqPvyrTPfQMI4LdMwfkY?usp=sharing

Public-health tool: https://nsr-models.onrender.com/

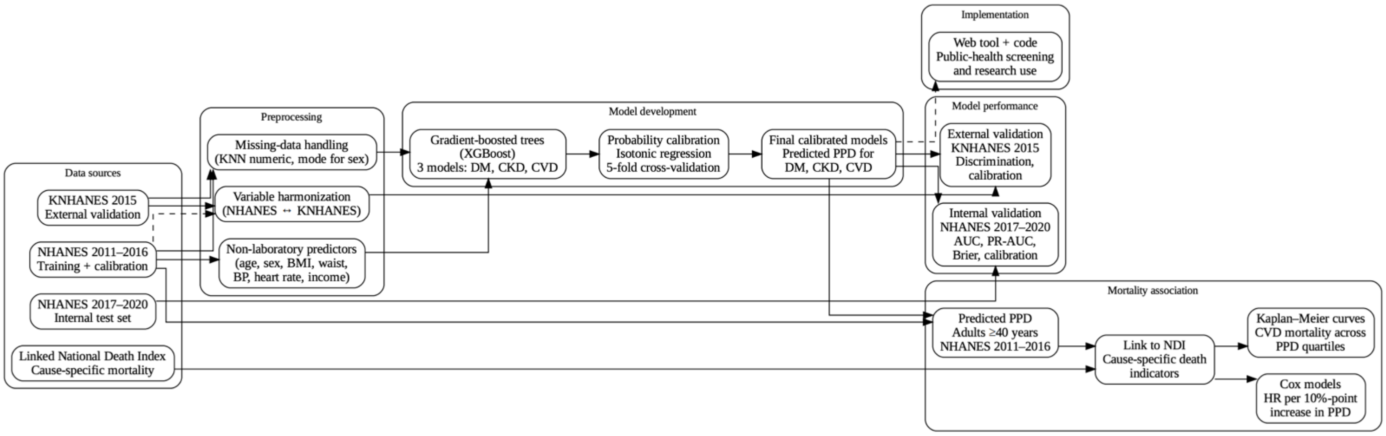


Figure S1: Model Creation and Usage Pipeline

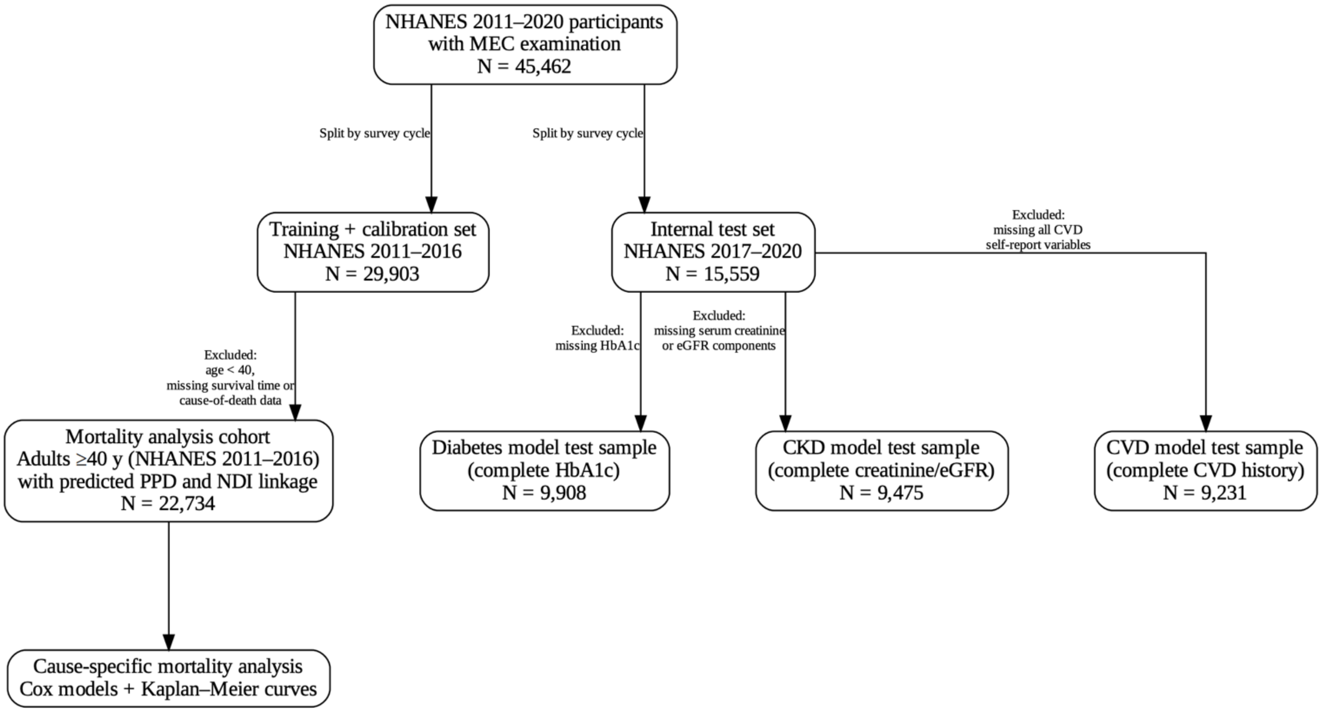


Figure S2. Flow of participants through the NHANES analytic pipeline.
Flow diagram showing the formation of the training, internal testing, and mortality-analysis cohorts derived from NHANES 2011–2020.

Table S1: Characteristics of the studied population according to sex [n (column%)].

| | | Total | Male | Female | P Value |
|---|---|---|---|---|---|
| | 1-19 | 11922 (26.2) | 6090 (27.4) | 5832 (25.1) | |
| | 20-29 | 6245 (13.7) | 3115 (14.0) | 3130 (13.5) | |
| | 30-39 | 5894 (13.0) | 2896 (13.0) | 2998 (12.9) | |
| | 40-49 | 6014 (13.2) | 2923 (13.2) | 3091 (13.3) | |
| | 50-59 | 6197 (13.6) | 3025 (13.6) | 3172 (13.6) | |
| | 60-69 | 4890 (10.8) | 2320 (10.4) | 2570 (11.1) | |
| | 70-79 | 2764 (6.1) | 1243 (5.6) | 1521 (6.5) | |
| Age | 80+ | 1539 (3.4) | 594 (2.7) | 945 (4.1) | <0.001 |
| | Mexican American | 4726 (10.4) | 2399 (10.8) | 2327 (10.0) | |
| Race | Other Hispanic | 3164 (7.0) | 1557 (7.0) | 1607 (6.9) | <0.001 |

| | | | | | |
|---|---|---|---|---|---|
| | Non-hispanic White | 27935 (61.4) | 13690 (61.7) | 14245 (61.3) | |
| | Non-hispanic Black | 5509 (12.1) | 2548 (11.5) | 2961 (12.7) | |
| | Non-hispanic Asian | 2436 (5.4) | 1155 (5.2) | 1281 (5.5) | |
| | Other or multiracial | 1692 (3.7) | 856 (3.9) | 836 (3.6) | |
| Education level - Adults 20+ | No high school education | 1732 (5.2) | 868 (5.4) | 864 (5.0) | |
| | Some high school education | 3202 (9.6) | 1668 (10.4) | 1534 (8.8) | |
| | High school graduate or GED | 7365 (22.0) | 3739 (23.2) | 3626 (20.8) | |
| | Some college or AA degree | 10760 (32.1) | 4787 (29.7) | 5973 (34.3) | |
| | College graduate or above | 10465 (31.2) | 5047 (31.3) | 5418 (31.1) | <0.001 |
| Smoking | Smoker | 10758 (42.5) | 6020 (49.3) | 4738 (36.1) | |
| | Non-smoker | 14564 (57.5) | 6181 (50.6) | 8383 (63.8) | <0.001 |

Table S2: Discrimination performance (AUC with 95% CI) across models

| **Model** | **Diabetes AUC (95% CI)** | **CKD AUC (95% CI)** | **CVD AUC (95% CI)** |
|---|---|---|---|
| Standard Only | 0.81 (0.80–0.82) | 0.89 (0.88–0.90) | 0.79 (0.78–0.80) |
| Diet + Standard | 0.83 (0.82–0.84) | 0.89 (0.88–0.90) | 0.79 (0.78–0.80) |
| Diet-Only (Avg) | 0.66 (0.64–0.67) | 0.67 (0.65–0.68) | 0.59 (0.57–0.60) |
| Diet-Only (Day 1) | 0.64 (0.62–0.65) | 0.66 (0.64–0.68) | 0.59 (0.57–0.60) |
| Diet-Only (Energy-Adjusted) | 0.66 (0.64–0.67) | 0.66 (0.64–0.68) | 0.58 (0.57–0.60) |

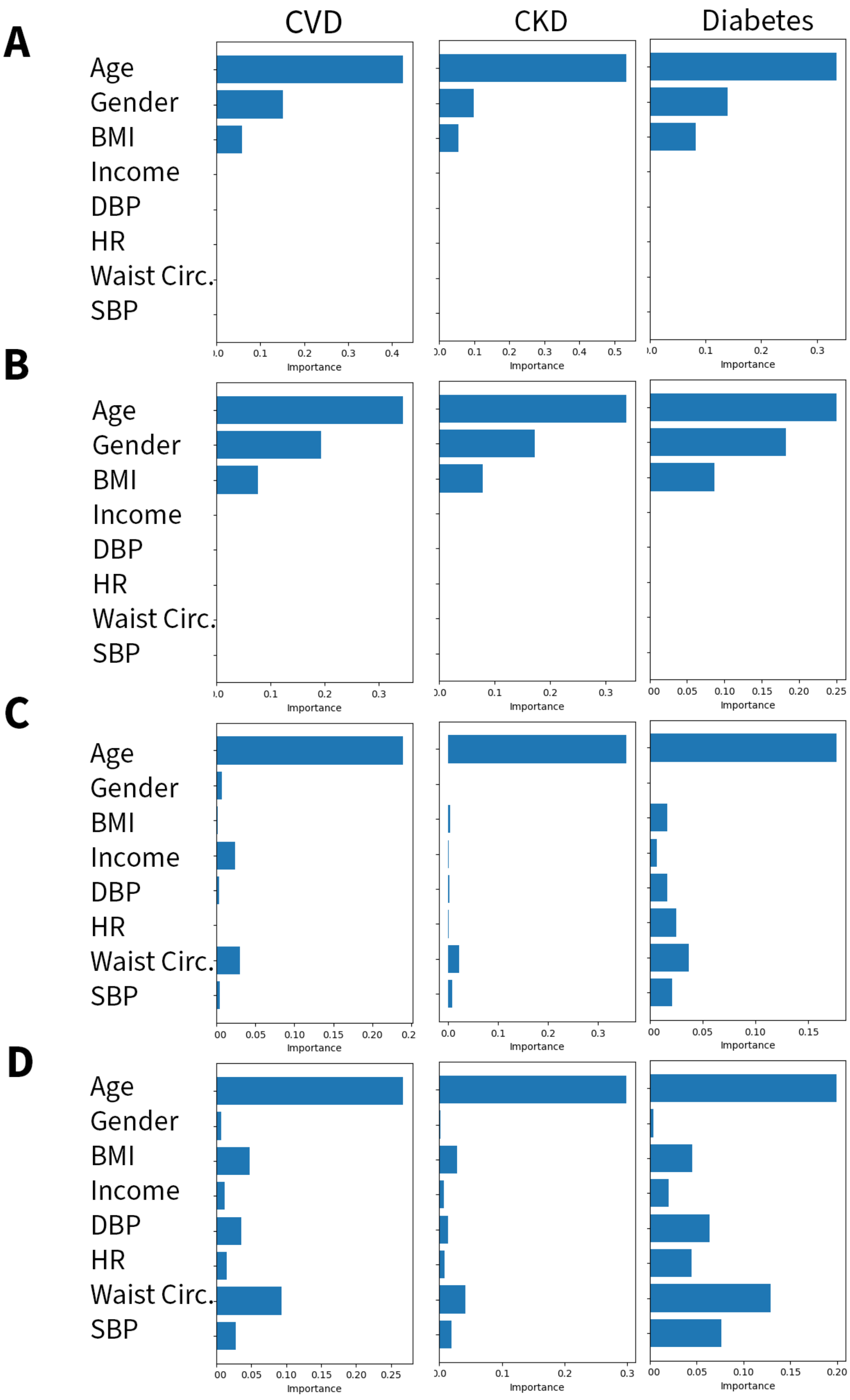

A
B
C
D
CVD
CKD
Diabetes
Age
Gender
BMI
Income
DBP
HR
Waist Circ.
SBP
Importance

Figure S3: CVD Feature Importance Plots. A) NHANES tree-based importance. B) KNHANES tree-based importance. C) NHANES permutation-based importance. D) KNHANES permutation-based importance.

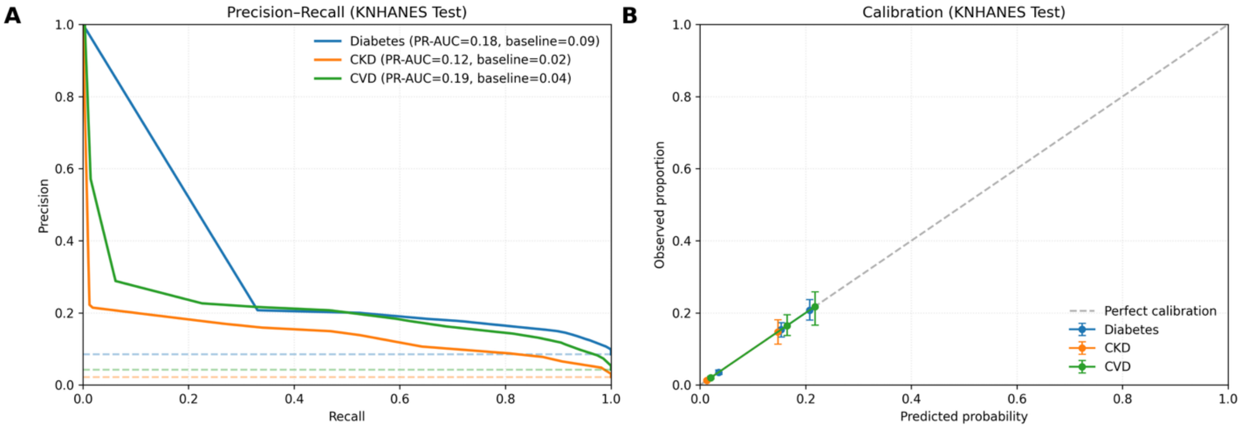


Figure S4. Precision-recall and calibration curves on KNHANES data. A) PR-AUC is reported and PR curves are compared to prevalence baselines, show in dotted lines. B) Model calibration is compared to perfect calibration across decile risk bins. Bins with less than 50 individuals are omitted. Error bars represent 95% CIs.

**Feature selection and importance sensitivity analyses**

We investigated the effect of excluding dietary recall data as predictors. Results are displayed in table S2. Adding 24- or 48-hour dietary recalls, with or without energy adjustment, to the standard predictor set did not improve discrimination for any outcome. Tree-based and permutation-based feature importance analyses showed that age, BMI, and waist circumference were dominant predictors of all 3 diseases (Fig. S2). We also evaluated whether basic tree ensembles with added low-value features (race, education, smoking, activity), simple imputation, and single-holdout calibration perform comparably to our feature-pruned, KNN-imputed, 5-fold cross validated, and boosted ensemble. These simpler models were not able to match the discrimination or calibration performance of our current models (Table S3). There was no difference in discrimination or calibration in KNN-imputed and mean/mode-imputed models, but every other change did individually improve AUC or Brier score when model changes were made one at a time. Finally, to characterize how the trained models behave across the observed NHANES covariate space, we created a series of partial dependence plots (PDPs) (Fig. S4).

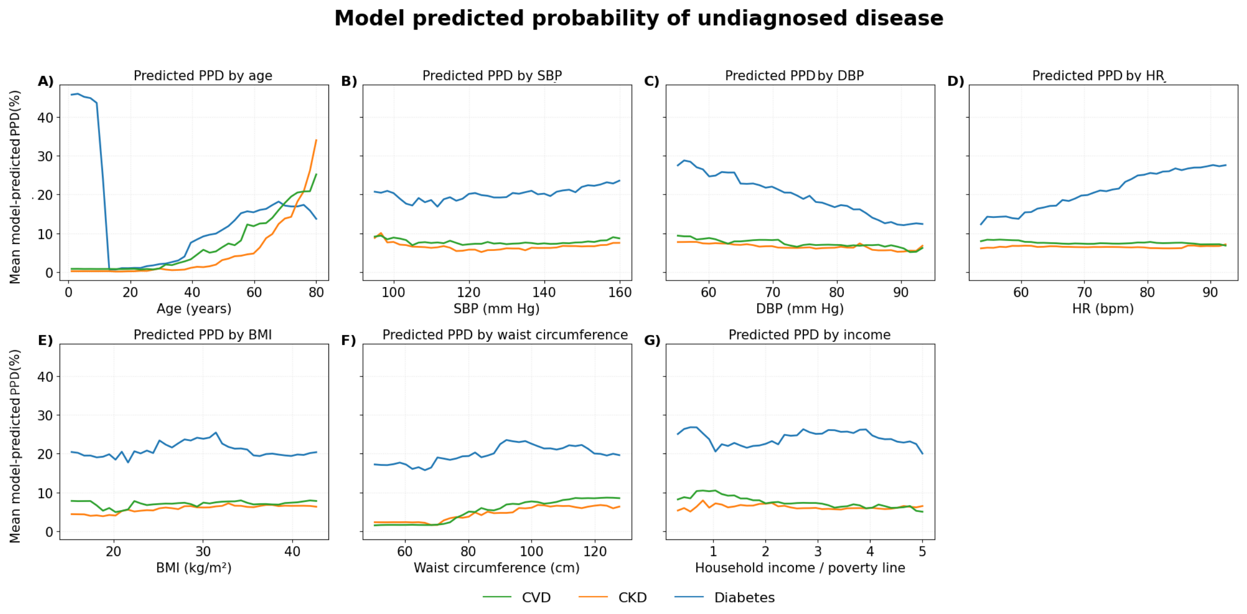


**Figure S5. PDP visualizations of model behavior**

Visualizations of model-predicted PPD as single predictors are varied. A) Age PDP. B) Systolic blood pressure PDP. C) Diastolic blood pressure PDP. D) Heart rate PDP. E) BMI PDP. F) Waist circumference PDP. G) Income PDP.

Table S3: Model comparisons

| Metric | Model | Diabetes | CKD | CVD |
|---|---|---|---|---|
| AUC (±95% CI) | Naïve | 0.806 ± 0.011 | 0.872 ± 0.012 | 0.772 ± 0.013 |
| | Main | 0.819 ± 0.010 | 0.895 ± 0.011 | 0.798 ± 0.012 |
| Brier (±95% CI) | Naïve | 0.0995 ± 0.0038 | 0.0571 ± 0.0033 | 0.1004 ± 0.0046 |
| | Main | 0.1001 ± 0.0042 | 0.0544 ± 0.0032 | 0.0966 ± 0.0042 |
| Calibration Slope | Naïve | 0.852 | 1.005 | 0.869 |
| | Main | 0.98 | 0.988 | 0.943 |

Naïve models are bagged tree models trained on all main predictors and the extra low-value predictors of race, education, activity, and smoking. These predictors were shown in our permutation-based feature importance to add approximately no value to predictive performance. Instead of using KNN imputation, the naïve model handled missingness by inserting training population means for continuous variables and modes for categorical variables. Instead of using isotonic recalibration with 5-fold cross validation, post-hoc recalibration of the naïve model was done with isotonic regression using a 5,000 person holdout from NHANES 2011-2016 that was not used for training. With this naïve recalibration method, calibration slopes were excessively affected by a few high-PPD predictions, so the top 1% of PPD subjects had to be excluded to achieve the moderate calibration seen. In all, the main model uses modern complex boosted tree modeling instead of bagged tree modeling, uses KNN instead of simple imputation, and uses 5-

fold cross-validation instead of holding out a calibration population en masse. Discrimination and calibration slope improved across all diseases, and calibration was able to be done without removing extreme PPD predictions. Alternative recalibration approaches, including Platt Scaling and Beta recalibration were also investigated, but showed decreases in Brier score compared to isotonic recalibration and were thus abandoned.

Table S4: Missingness description

| **Variable name** | **Train missing n (%)** | **Test missing n (%)** | **KNHANES missing n (%)** | **Likely Reasons for Missingness** |
|---|---|---|---|---|
| AgeYears | 0 (0.0%) | 0 (0.0%) | 0 (0.0%) | - |
| Gender | 0 (0.0%) | 0 (0.0%) | 0 (0.0%) | - |
| FamIncome_to_poverty_ratio | 2,677 (9.0%) | 2,201 (14.2%) | 52 (0.7%) | Respondent refusal or income nondisclosure; not collected for participants <15; missing in low-income household subsamples. |
| bmi | 3,490 (11.7%) | 2,422 (15.6%) | 421 (5.7%) | Missing when physical exam not completed or participant declined anthropometry. Commonly missing in very young children. |
| waist_circumference | 4,725 (15.8%) | 2,985 (19.2%) | 414 (5.6%) | Not measured for children <2; participants refusing waist measurement; incomplete MEC exams. |
| avg_ systolic | 7,961 (26.6%) | 5,206 (33.5%) | 1,104 (15.0%) | Missing when blood pressure could not be measured (equipment error, participant refusal, age <8 years). |
| avg_ diastolic | 7,961 (26.6%) | 5,206 (33.5%) | 1,104 (15.0%) | Same reasons as systolic BP; diastolic missingness tracks systolic exactly. |
| avg_HR | 2,074 (6.9%) | 6,087 (39.1%) | 1,569 (21.3%) | Resting pulse not always recorded; sometimes measured only once or skipped due to movement or irregularity readings. |
| binary_ diabetes | 16,820 (56.3%) | 5,651 (36.3%) | 1,556 (21.1%) | Requires lab measures (HbA1c); missing when blood draw not completed or for children (<12). |
| binary_ CKD | 11,119 (37.2%) | 6,084 (39.1%) | 0 (0.0%) | Requires serum creatinine for eGFR; missing when lab module not completed. Nearly complete in KNHANES due to high lab compliance. |
| binary_cvd | 12,859 (43.0%) | 6,328 (40.7%) | 921 (12.5%) | Based on self-report of physician-diagnosed heart disease; not asked to |

| | | | | |
|---|---|---|---|---|
| | | | | children; extensive skip patterns; refusal/missing interviews. |

Table S5. Variable dictionary

| Variable | Description | NHANES Variable Name(s) | KNHANES Variable Name(s) | Coding (post-transformations) |
|---|---|---|---|---|
| AgeYears | Participant age in years at time of exam/interview | RIDAGEYR | age / AGE | Continuous (years) |
| Gender | Participant sex | RIAGENDR | sex / SEX | Categorical: typically Male, Female |
| FamIncome_to_poverty_ratio | Family income relative to the U.S. (or national) poverty threshold | INDFMPIR | inc_poverty_ratio (constructed from income quantiles) | Continuous (ratio) |
| bmi | Body Mass Index | BMXBMI | BMI | Continuous (kg/m²) |
| waist_circumference | Waist circumference | BMXWAIST | waist_circumference / WC | Continuous (cm) |
| avg_systolic | Average systolic blood pressure | BPXSY1, BPXSY2, BPXSY3 (averaged) | SBP1, SBP2, SBP3 (averaged) | Continuous (mmHg) |
| avg_diastolic | Average diastolic blood pressure | BPXDI1, BPXDI2, BPXDI3 (averaged) | DBP1, DBP2, DBP3 (averaged) | Continuous (mmHg) |
| avg_HR | Resting heart rate | BPXPLS1, BPXPLS2 (averaged) | pulse, PULSE | Continuous (beats per minute) |
| activity_level | Overall physical activity category based on WHO guidelines | PAQ* series (e.g., PAQ605, PAQ620) + PAD* duration items | Not used | Categorical: Low, Moderate, High |
| binary_diabetes | Diabetes status based on glycemic laboratory values | LBXGH (HbA1c) | HE_L_HBA1C or similar (HbA1c) | Binary (0/1) |

| | | | | |
|---|---|---|---|---|
| binary_CKD | Chronic kidney disease status | LBXSCR (serum creatinine) | creatinine / HE_creatinine | Binary (0/1) |
| binary_cvd | Cardiovascular disease status based on self-reported history | MCQ160B, MCQ160C, MCQ160D, MCQ160E, MCQ160F | doctor_MI, doctor_angina | Binary (0/1). **MISMATCH.** |

Gradient-boosted tree models were implemented using XGBoost 3.1.2 and scikit-learn 1.6.1. For each outcome, we trained an XGBClassifier with 300 boosting rounds (n_estimators=300), a maximum tree depth of 4 (max_depth=4), a learning rate of 0.05, and subsampling and column-subsampling rates of 0.8 (subsample=0.8, colsample_bytree=0.8). The loss function was binary logistic (objective="binary:logistic") with log-loss evaluation (eval_metric="logloss"), and trees were constructed using histogram-based splitting (tree_method="hist"). To account for class imbalance, we applied outcome-specific scale_pos_weight values computed from the ratio of negative to positive cases in the training data. All models used random_state=42 and parallel computation (n_jobs=-1), with remaining XGBoost parameters left at their defaults.

Predictor preprocessing was performed using scikit-learn Pipelines. Numeric variables were imputed using KNNImputer (n_neighbors=5, weights="distance"), while the categorical sex variable was imputed using SimpleImputer(strategy="most_frequent") followed by one-hot encoding with handle_unknown="ignore". Model probabilities were calibrated using isotonic regression. NHANES models were calibrated via 5-fold isotonic cross-validation (CalibratedClassifierCV with method="isotonic", cv=5). For external validation, KNHANES models were recalibrated by fitting a single isotonic regression mapping to predictions from the NHANES-trained model. All remaining preprocessing and calibration hyperparameters were left at their scikit-learn defaults.

Table S6. Model hyperparameters

| **n_estimators** | **max_depth** | **Learning_rate** | **Subsample** | **Colsample_bytree** | **tree_method** | **DM scale_pos_weight** | **CKD scale_pos_weight** | **CVD scale_pos_weight** |
|---|---|---|---|---|---|---|---|---|
| 300 | 4 | 0.05 | 0.8 | 0.8 | hist | 8.23 | 13.12 | 8.41 |
